# Clinical accuracy of SARS-CoV-2 rapid antigen testing in screening children and adolescents in comparison to RT-qPCR, November 2020 to September 2022

**DOI:** 10.1101/2022.11.07.22281809

**Authors:** Manuel Krone, Isabell Wagenhäuser, Kerstin Knies, Daniela Hofmann, Geraldine Engels, Regina Taurines, Miriam McDonogh, Sven Flemming, Thomas Meyer, Hartmut Böhm, Agmal Scherzad, Michael Eisenmann, Vera Rauschenberger, Alexander Gabel, Nils Petri, Julia Reusch, Johannes Forster, Benedikt Weißbrich, Lars Dölken, Oliver Kurzai, Ulrich Vogel, Christoph Härtel, Johannes Liese, Oliver Andres

**Affiliations:** Infection Control and Antimicrobial Stewardship Unit, University Hospital Wuerzburg, Wuerzburg, Germany; Institute for Hygiene and Microbiology, University of Wuerzburg, Wuerzburg, Germany; Department of Internal Medicine I, University Hospital Wuerzburg, Wuerzburg, Germany; Institute for Virology and Immunobiology, University of Wuerzburg, Wuerzburg, Germany; Department of Paediatrics, University Hospital Wuerzburg, Wuerzburg, Germany; Department of Child and Adolescent Psychiatry, Psychosomatics and Psychotherapy, University Hospital Wuerzburg, Wuerzburg, Germany; Department of Orthopaedic Trauma, Hand, Plastic and Reconstructive Surgery, University Hospital Wuerzburg, Wuerzburg, Germany; Department of General, Visceral, Transplantation, Vascular and Paediatric Surgery, University Hospital Wuerzburg, Wuerzburg, Germany; Department of Oral and Maxillofacial Surgery, University Hospital Wuerzburg, Wuerzburg, Germany; Department of Otorhinolaryngology, Plastic, Aesthetic and Reconstructive Head and Neck Surgery, University Hospital Wuerzburg, Wuerzburg, Germany; Leibniz Institute for Natural Product Research and Infection Biology – Hans-Knoell-Institute, Jena, Germany

**Keywords:** SARS-CoV-2 antigen rapid diagnostic test, PCR, clinical performance evaluation, paediatric, children

## Abstract

**Background:** Rapid antigen detection tests (RDT) are an easily accessible, feasible, inexpensive, and point-of-care method in SARS-CoV-2 diagnostics – established in adults as well as in children and adolescents. Despite this, large-scale data of clinical performance in the paediatric population especially regarding the influence of SARS-CoV-2 virus variants of concern (VOC) and COVID-19 vaccination on test accuracy is rare.

**Methods:** This single-centre prospective diagnostic study evaluates three RDT (NADAL®, Panbio™, MEDsan®) in comparison to quantitative reverse transcription polymerase chain reaction (RT-qPCR). 9,760 oropharyngeal screening samples regarding SARS-CoV-2 VOC and COVID-19 vaccination in paediatric hospitalised patients aged younger than 18 years were enrolled.

**Findings:** RDT sensitivity was 44·7% (157/351, 95% CI 39·6%–50·0%) compared to the reference standard RT-qPCR, specificity 99·8% (9,392/9,409, 95% CI 99·7%–99·9%). Most SARS-CoV-2 infections considered were caused by Omicron VOC. Diagnostic accuracy of RDT depended on specimen containing viral load with a decreasing RDT sensitivity by descending viral load, corresponding with a significantly impaired sensitivity in asymptomatic children. A sensitivity of 71·0% was obtained for a viral load higher than 10^6^ SARS-CoV-2 RNA copies per ml suggested as infectivity threshold. No significant differences in RDT sensitivity could be observed regarding gender, symptoms, COVID-19 vaccination status, and VOC.

**Interpretation:** In a paediatric population, RDT have proven to reliably detect potentially highly infectious patients with a viral load of at least 10^6^ SARS-CoV-2 RNA copies per ml. Due to the low sensitivity in asymptomatic individuals, the usefulness of RDT seems limited in large-scale SARS-CoV-2 screening programs.

**Funding:** Federal Ministry for Education and Science (BMBF), Free State of Bavaria

## Introduction

The COVID-19 pandemic and its wide reaching consequences has been heading international efforts since early 2020. To establish effective and rapid SARS-CoV-2 diagnostics is one of the keys mitigating COVID-19 pandemic dynamics. The earliest possible detection of SARS-CoV-2 infected individuals is the central pillar of a hospital’s screening strategy avoiding nosocomial outbreaks of SARS-CoV-2 infections and transmission clusters.^1,2^

The reference standard for SARS-CoV-2 detection is defined as a quantitative reverse transcription polymerase chain reaction (RT-qPCR).^3^ However, usability and availability are limited due to the need of technical and laboratory infrastructure, high cost, and longer analysis times. The enormous acceleration of the global SARS-CoV-2 incidence acceleration due to dissemination of SARS-CoV-2 spike protein variants of concern (VOC), especially the Delta and Omicron VOC, highlighted the limited capacities of RT-qPCR and the need of an alternative diagnostic strategy.^4,5^

Over the past two years, more than 600 different SARS-CoV-2 rapid antigen detection tests (RDT) became available promising cost-effective, affordable rapid point-of-care diagnostics as well as self-testing options for broad screening use.^6,7^ Despite this, the available evidence focuses mainly on an adult study population. A large-scale, real-life analysis of RDT performance among children and adolescents including COVID-19 vaccination status and SARS-CoV-2 VOC is still missing.^8-18^

The aim of this prospective performance evaluation study is to evaluate the clinical performance of RDT compared to RT-qPCR as screening test strategy for hospitalised children and adolescents under the age of 18 years. The analysis includes a standardised viral load determination as reference with the focus on SARS-CoV-2 VOC dependence, COVID-19 vaccination status, and specificity in broad screening use.

**Research in context**

*Question*

PubMED and medRxiv were screened for evidence including the terms “COVID-19”, “COVID”, “SARS-CoV-2”, “coronavirus”, “virus variants of concern”, “Omicron”, “Delta”, “Alpha”, “B.1.617.2”, “B.1.1.529”, “B.1.1.7”, “children”, “paediatric”, “adolescents” as well as “antigen detection”, “rapid antigen test”, “point-of-care test”, “diagnostics”, in title or abstract, published between 1^st^ January of 2020 and 30^th^ September of 2022.

A large and important cohort of individuals tested by RDT in clinical as well as in broad screening use are children and adolescents. Despite the possible grave impact of the COVID-19 pandemic on childhood and adolescence regarding physical and mental health, previous examinations focused predominantly on adults. Comparable clinical RDT performance data from a large-scale paediatric cohort is lacking.

Further, transferable, real-life evidence considering the potential influence of VOC on RDT performance for a paediatric cohort is still limited while laboratory studies report a restricted RDT sensitivity to detect Omicron VOC. Clinical data in adults is ambiguous. Although COVID-19 vaccinations have proven as an important SARS-CoV-2 prevention strategy in school age children and adolescents, the influence on RDT performance is still unclear for this cohort.^14^ From the paediatric perspective, RDT performance including the influence of VOC, age, and COVID-19 vaccination status has to be investigated.

*Added value of this study*

The study provides a large-scale analysis based on the daily clinical screening routine of a tertiary care hospital and enables transferability of data to the general public. This is the first paediatric RDT performance assessment which considers COVID-19 vaccination status and VOC as potential influencing factors on the RDT performance.

Among children and adolescents, RDT sensitivity compared to RT-qPCR was 44·7%, specificity 99·8%. Based on a multiple regression analysis, RDT sensitivity depended significantly on viral load, age, and days since symptom onset and is not statistically significant influenced by gender, symptoms, COVID-19 vaccination status, and VOC. For highly infectious children with a viral load of at least 10^6^ SARS-CoV-2 RNA copies per ml suggested as infectivity threshold sensitivity was 71·0%.

*Implications of all the available evidence*

For paediatric screening usage, RDT have proven be a reliable diagnostic tool for SARS-CoV-2 infected children presenting high viral loads beyond 10^6^ SARS-CoV-2 RNA copies per ml. Due to the low sensitivity in asymptomatic individuals, the usefulness of RDT seems limited in large-scale SARS-CoV-2 screening programs.

## Methods

### Study setting

From the 12^th^ of November 2020 to the 30^th^ of September 2022, the clinical RDT performance evaluation took place in a tertiary care hospital in Bavaria, Germany. The data collection period covers the second up to the beginning of the seventh wave of COVID-19 pandemic in Germany, driven predominantly by wild-type SARS-CoV-2, Alpha VOC, Delta VOC, as well as the Omicron VOC with its sublineages BA.1, BA.2, BA.4 and BA.5.^4,19^ During the study period, the weekly average SARS-CoV-2 incidence in the Federal State of Bavaria was 457 per 100,000 inhabitants in the general population and 554 per 100,000 in inhabitants aged younger than 16 years. Separate SARS-CoV-2 incidence data for adolescents aged 16 to 18 years was not available by the federal statistical office.^4^

This study is a following paediatric specific sub-study of two former RDT performance assessments from November 2020 up to February 2021, the first of which included 1,034 RDT on paediatric patients in a total of 5,068 RDT,^20^ as wells as its follow-up study up to January 2022 with 35,479 enrolled specimens including 5,623 from children and adolescents.^18^

### Test enrolment

As part of the hospital’s set of measures for prevention and reduction of intrahospital SARS-CoV-2 dissemination, a tandem RDT and RT-qPCR test concept was established in all patients undergoing hospitalisation or procedures. In the paediatric department and other critical areas, such as the emergency department this strategy was performed for the duration of the entire study period. Other departments which also treat paediatric patients, RDT use combined with RT-qPCR was restricted for all patients on admission to pandemic intervals of a high SARS-CoV-2 incidence (1^st^ of February to 30^th^ of June 2021, and 4^th^ of November 2021 to 30^th^ of September 2022). This approach met the rationale of the parallel RDT/RT-qPCR screening to enable the rapid isolation of infectious children and adolescents in case of RDT positivity before the completion of the RT-qPCR in the setting of hospitalisation.

Only the first RDT per day per person was considered for data analysis. In participants met the inclusion criteria on multiple days during the study period, only the first daily RDT result was considered. SARS-CoV-2 convalescent individuals with subsequent deisolation were excluded in view of a potential persistent RT-qPCR positivity without relation to a risk of viral spread.^20^

### Antigen rapid diagnostic tests (RDT)

Three different RDT were selected due to manufacturers’ performance data and availability at the beginning of the study.^20^ All used RDT target SARS-CoV-2 nucleoprotein antigen.

I. NADAL® COVID-19 Ag Test (Nal von Minden GmbH, Regensburg, Germany)
II. PANBIO™ COVID-19 Ag Rapid Test (Abbott Laboratories, Abbott Park IL, USA)
III. MEDsan® SARS-CoV-2 Antigen Rapid Test (MEDsan GmbH, Hamburg, Germany)

The chosen RDT were continuously applied during the entire study period, having been randomly distributed to the hospital departments due to the local infrastructure and RDT availability.

Invalid RDT results were considered as a repetition of the RDT. Because only the first RDT per day per person was included, results of repeated RDT were excluded.

RDT and RT-qPCR specimens were obtained in successive paired oropharyngeal swabs by trained medical professionals in accordance with manufacturers’ instructions.

### Quantitative reverse transcription polymerase chain reaction (RT-qPCR)

RT-qPCR was performed on oropharyngeal swabs using different CE marked analytic devices and PCR kits in the virological diagnostic laboratories according with manufacturers’ instructions as described earlier.^20^

From the 3^rd^ of February 2021 to the 19^th^ of January 2022 a spike protein variant specific PCR was performed on all new RT-qPCR positive samples. As the proportion of the Omicron VOC exceeded 95% of the entire SARS-CoV-2 cases in Germany, VOC specification was interrupted due to limited diagnostic capacities.^20^ If VOC determination failed in samples with low viral loads or if the sample was collected outside the VOC determination period, samples were assigned to the dominating VOC at that point of time in Germany. This VOC was defined as the one detected in more than 90% of all SARS-CoV-2 positive samples in that calendar week in Germany as described earlier.^19,20^

Calculation of viral loads starting from cycle threshold (Ct) values based on reference standards as well as molecular VOC determination were defined previously as well.^18,20^

### Data collection

Documented RDT and RT-qPCR results combined with demographic data, SARS-CoV-2 infection related symptomatology were extracted from the local hospital information system (HIS) SAP ERP 6.0 (SAP, Walldorf, Germany), information on COVID-19 vaccination status from the hospital’s standardised COVID-19 clinical admission questionnaire. All RDT performed before the official EMA (European Medicines Agency) vaccination authorisation date of BNT162b2mRNA (Comirnaty, BioNTech/Pfizer, Mainz/Germany, New York/USA) for children and adolescents without information on COVID-19 vaccination status were classified as performed on COVID-19 unvaccinated individuals: for participants aged 16 and 17 years the entire of RDT before the 12^th^ of December 2020, for 12 to 15 years aged before the 31^st^ of May 2021 and for 5 to 11 years aged before the 26^th^ of November 2021.^21^

Included individuals were classified in three categories based on COVID-19 related symptoms following the COVID-19 case definitions of the CDC and the ECDC:^22^

A. asymptomatic
B. atypical symptomatic possibly caused directly or indirectly by SARS-CoV-2 (e.g. deterioration of general condition, falls, diarrhoea, or seizures)
C. typical symptomatic (e.g. fever, dry cough, shortness of breath, new anosmia or ageusia)

The following age categories were set for an age-stratified analysis of RDT performance, viral load, COVID-19 symptomatology, and days since symptom onset: first year of life, 1 to 5 years, 6 to 11 year and 12 to 17 years.

### Statistics

Data analysis was performed using GraphPad Prism 9.4.1 (GraphPad Software, San Diego CA, USA) and R (version 4.1.3).^23^

Confidence intervals were calculated with the Wilson/Brown method,^24^ statistical significance levels were calculated using Fisher’s exact test (comparison of manufacturers, VOC, vaccination status and symptomatology and age category specific sensitivity), and Mann Whitney U-test (comparison of viral loads and age categories). The two-tailed significance level α was set to 0·05.

A logistic lasso regression analysis was performed to identify factors that are associated with the result of RDT to confirm a SARS-CoV-2 infection. The regression model included the factors viral load, age, symptoms, gender, COVID-19 vaccination status, and being infected by the Omicron VOC. Using a tenfold cross-validation procedure, the model parameters of the lasso regression model were estimated, and the model having the lowest mean squared error (MSE) of ∼0·89 was chosen.

### Ethical approval

The Ethics committee of the University of Wuerzburg considered the study protocol and waived the need to formally apply for ethical clearance due to the study design (File 20220602 01).

## Results

### Test enrolment

During the study period from 12^th^ of November 2020 to 30^th^ of September 2022, 9,939 RDT with parallel RT-qPCR were performed. 160 tests were excluded due to multiple RDT performance per day per patient. Twelve RDT performed on individuals recently deisolated after a SARS-CoV-2 infection with a positive RT-qPCR result were not considered. Seven performed RDT were invalid (by virtue of lacking control or interfering lines: one NADAL®, two Panbio™, four MEDsan®) and consequently debarred.

9,760 RDT results on 7,472 children and adolescents aged 0 to 17 years were enrolled and included into data analysis. NADAL® was used in 740 (7·6%), PANBIO™ in 3,960 (40·6%), and MEDsan® in 5,060 (51·8%) RDT (*Figure 1*).

**Figure 1:**
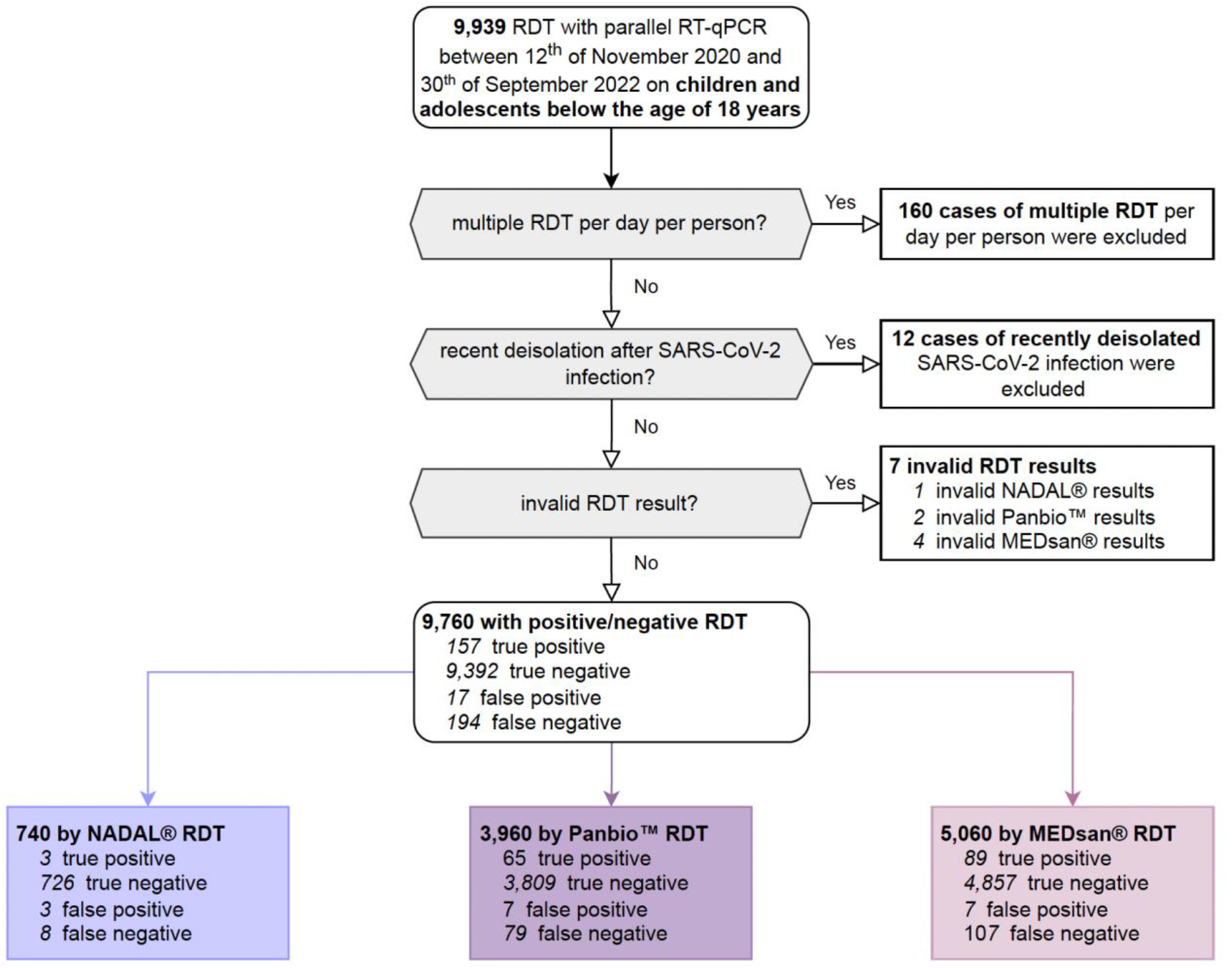
Enrolment of antigen rapid diagnostic test (RDT) results RDT: Antigen rapid diagnostic test. RT-qPCR: Quantitative reverse transcription polymerase chain reaction.

### Study population

The age distribution of the cohort as presented in *Figure 2* is characterised by a peak in children in the first year of life and a nadir in 9-year-old children (median age: 5 years, IQR: 1-14 years). As majority 6,227 (63·8%) of the RDT were performed in the Department of Paediatrics. A further 1,419 (14·5%) RDT were conducted in both local surgical departments, and 1,127 (11·5%) in the Department of Child and Adolescent Psychiatry, Psychosomatics and Psychotherapy (CAP). The remaining 987 (10·1%) RDT were distributed over other departments of the hospital (*Figure 2*).

**Figure 2:**
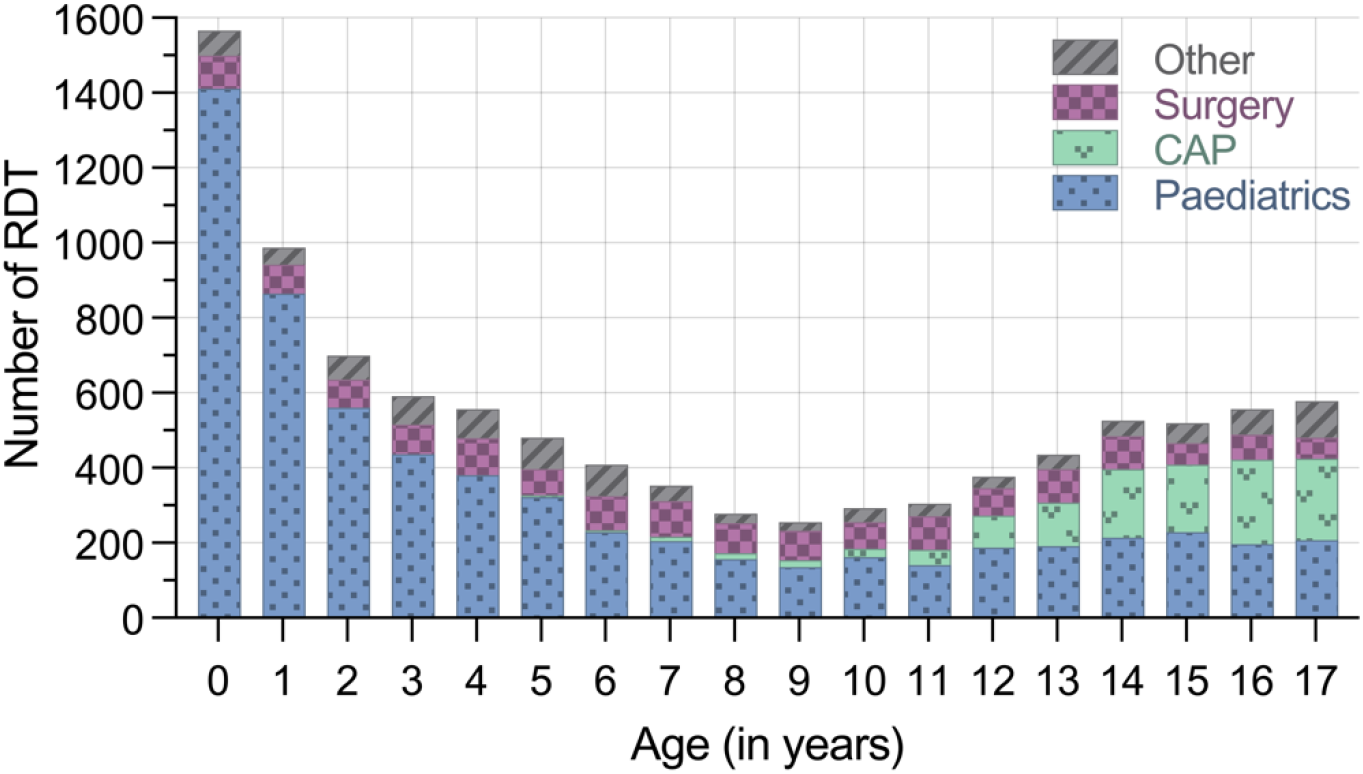
Distribution of patients regarding age and testing department Following the initial peak in the first year of life the RDT were distributed relatively uniformly from the age of 2 up to 17 years (n=9,760). While enrolled RDT in children were predominantly carried out in the Department of Paediatrics, the majority of RDT in adolescents aged 16 to 17 years was performed at the Department of Child and Adolescent Psychiatry, Psychosomatics and Psychotherapy (CAP) representing the demography of underage patients admitted to the hospital. RDT: Antigen rapid diagnostic test CAP: Child and Adolescent Psychiatry, Psychosomatics and Psychotherapy

5,195 (53·2%) RDT were performed on patients assigning themselves to male gender, 4,564 (46·8%) on patients assigned to female gender. One RDT was performed on a patient of diverse gender.

### RDT performance compared to RT-qPCR

Out of 9,760 enrolled RDT/RT-qPCR tandems, 351 samples tested positive for SARS-CoV-2 by RT-qPCR representing a SARS-CoV-2 prevalence of 3·6%. 157 of the RT-qPCR positive cases had a positive RDT result (true positive), 194 a negative RDT result (false negative). The remaining 9,409 samples were RT-qPCR negative whereof 9,392 tested RDT negative (true negative) and 17 RDT positive (false positive; *Figure 2*). Two of the seven invalid RDT had a corresponding SARS-CoV-2 positive RT-qPCR.

The overall sensitivity was 44·7% (157/351, 95% CI 39·6%–50·0%, *Figure 3A*), overall specificity 99·8% (9,392/9,409, 95% CI 99·7%–99·9%, *Figure 3B*). The positive predictive value (PPV) was obtained as 90·2% (157/174, 95% CI 84·9%–93·8%), the negative predictive value (NPV) as 98·0% (9,392/9,586, 95% CI 97·7–98·2%).

**Figure 3:**
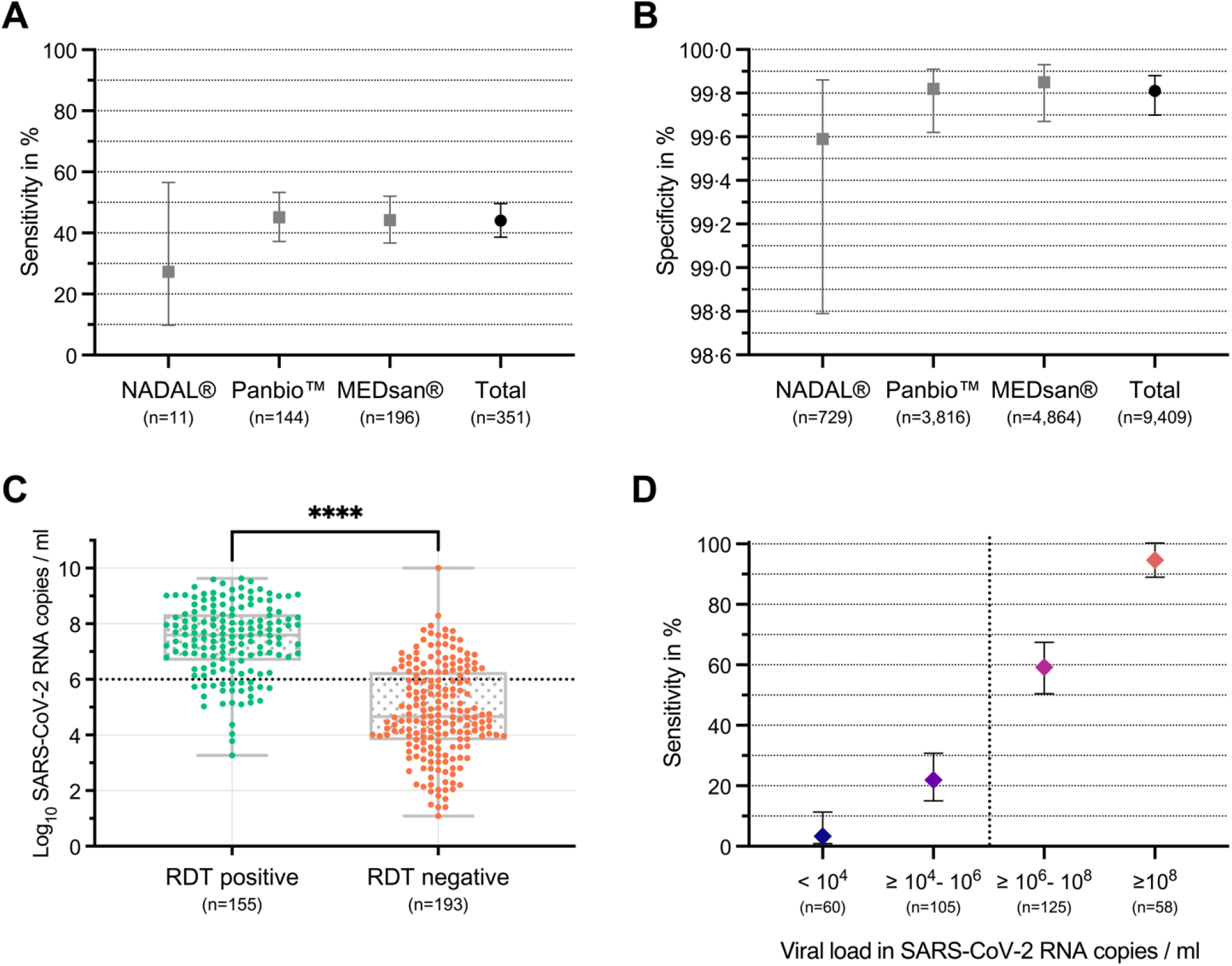
Analytical performance of 9,760 RDT performed in underage individuals compared to RT-qPCR **Figure 3A and 3B**: Sensitivity and specificity of RDT compared to RT-qPCR as reference diagnostics in total and separated by RDT manufacturer (nal von minden NADAL®, Abbott Panbio™, MEDsan®), n=9,760. **Figure 3C** displays the logarithmised viral load in SARS-CoV-2 RNA copies/ml, separated by RDT results. No viral load could be obtained for three RDT (one RDT with a negative, two with a positive result), n=348. The viral load of specimen with positive RDT results (true positive) exceeded statistically significant the viral load of RT-qPCR positive samples with negative RDT result (false negative). **Figure 3D**: RDT sensitivity depending on specimen containing viral load, calculated based on standardised Ct values, n=348. The viral load threshold of 10^6^ SARS-CoV-2 RNA copies/ml, suggested as infectivity threshold, is added as horizontal dotted line to Figure 3C and vertical dotted line to Figure 3D.^25^ n: Number of enrolled RDT per group RDT: Antigen rapid diagnostic test RT-qPCR: Quantitative reverse transcription polymerase chain reaction ****: p<0·0001

### Comparison of RDT manufacturers

The differences in sensitivity (all pairwise comparisons using Fisher’s exact test p>0·34, *Figure 3A*) and specificity (all p>0·13, *Figure 3B*) were not significant.

### Influencing factors on RDT performance

In the logistic lasso regression analysis, the factors viral load and Omicron VOC infection showed associations influencing the result of RDT (*Figure 4*). Based on the logistic regression, the concentration of viral load significantly increases the odds of having a positive test (p<0·0001) while the independent negative influence of Omicron VOC on the RDT result was not significant (p=0·12).

**Figure 4:**
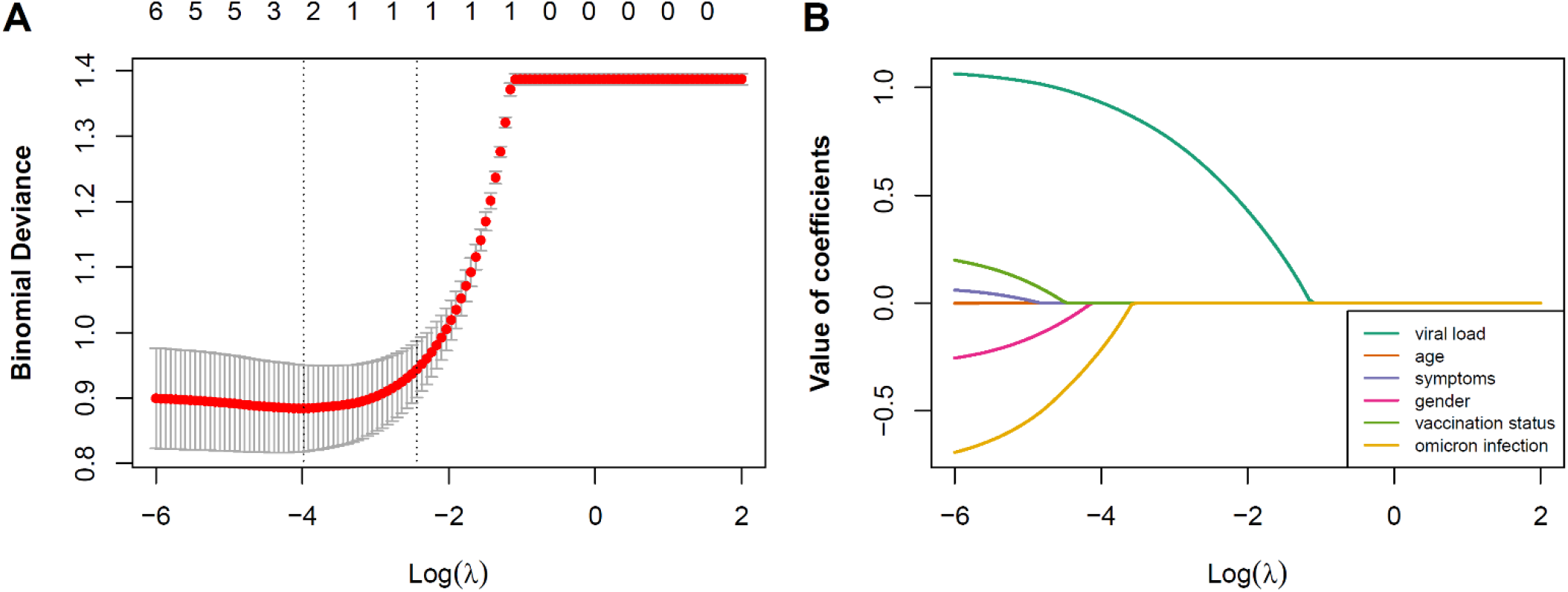
Lasso regression for detection of influencing factors associated to RDT sensitivity **Figure 4A**: Tenfold cross-validation procedure to determine optimal lambda parameter based on minimal mean-squared error. **Figure 4B**: Illustrating the shrinkage of coefficients (factors) towards zero with increasing lambda values.

### Influence of viral load on RDT performance

For 348 out of the 351 SARS-CoV-2 positive samples, the viral load was determined. No viral load could be obtained for three RDT (one RDT with a negative, two with a positive result).

A median viral load of 1·8×10^6^ (IQR: 2·7×10^4^-3·8×10^7^) SARS-CoV-2 RNA copies per ml was detected among all RT-qPCR positive specimen. Significantly higher median viral loads were obtained for RDT positive samples (155/348, median: 4·0×10^7^, IQR: 4·6×10^6^-2·2×10^8^ SARS-CoV-2 RNA copies per ml) than for RDT negative samples (193/348, median: 4·6×10^4^, IQR: 6·4×10^3^-1·9×10^6^ SARS-CoV-2 RNA copies per ml, p<0·0001, Mann-Whitney U test, *Figure 3C*).

RDT sensitivity increased significantly by viral load: for specimen containing an estimated viral load lower than 10^4^ SARS-CoV-2 RNA copies per ml, RDT sensitivity was 3·3% (2/60, 95% CI 0·9%–11·4%), increasing for higher viral load categories up to 96·6% (56/58, 95% CI 88·3%–99·1%) for samples containing at least 10^8^ SARS-CoV-2 RNA copies per ml. Considering the viral load threshold of 10^6^ SARS-CoV-2 RNA copies per ml, suggested as SARS-CoV-2 infectivity threshold,^25^ RDT sensitivity was 71·0% (130/183, 95% CI 64·1%–77·1%) for samples containing viral loads above this limit (*Figure 3D*).

### Influence of symptomatology on viral load and RDT performance

Among the 351 RT-qPCR confirmed SARS-CoV-2 positive children and adolescents, information on COVID-19 infection related symptoms was available in 339 cases: 78 (23·0%) presented as asymptomatic, 32 (9·4%) were categorised as atypically, and 229 (67·6%) as typically COVID-19 symptomatic.^26^

In the subgroup of asymptomatic individuals RDT sensitivity was 20·5% (16/78, 95% CI 13·0%–30·8%), among atypically symptomatic 50.0% (16/32, 95% CI 33·6%–66·4%), and 53·3% (122/229, 95% CI 46·8%–59·6%) among the typically symptomatic children and adolescents. The median viral load was 2·6×10^4^ (IQR: 2·8×10^3^-5·5×10^5^) SARS-CoV-2 RNA copies per ml among asymptomatic, 2·5×10^6^ (IQR: 8·6×10^4^-9·7×10^7^) SARS-CoV-2 RNA copies per ml among atypically symptomatic, and 6·1×10^6^ (IQR: 1·9×10^5^-6·4×10^7^) SARS-CoV-2 RNA copies per ml among typically symptomatic children and adolescents.

Asymptomatic children and adolescents presented a median viral load of. RDT sensitivity was significantly reduced in asymptomatic compared to the symptomatic children and adolescents (p=0·0022, Fisher’s exact test, *Figure 5D*).

**Figure 5:**
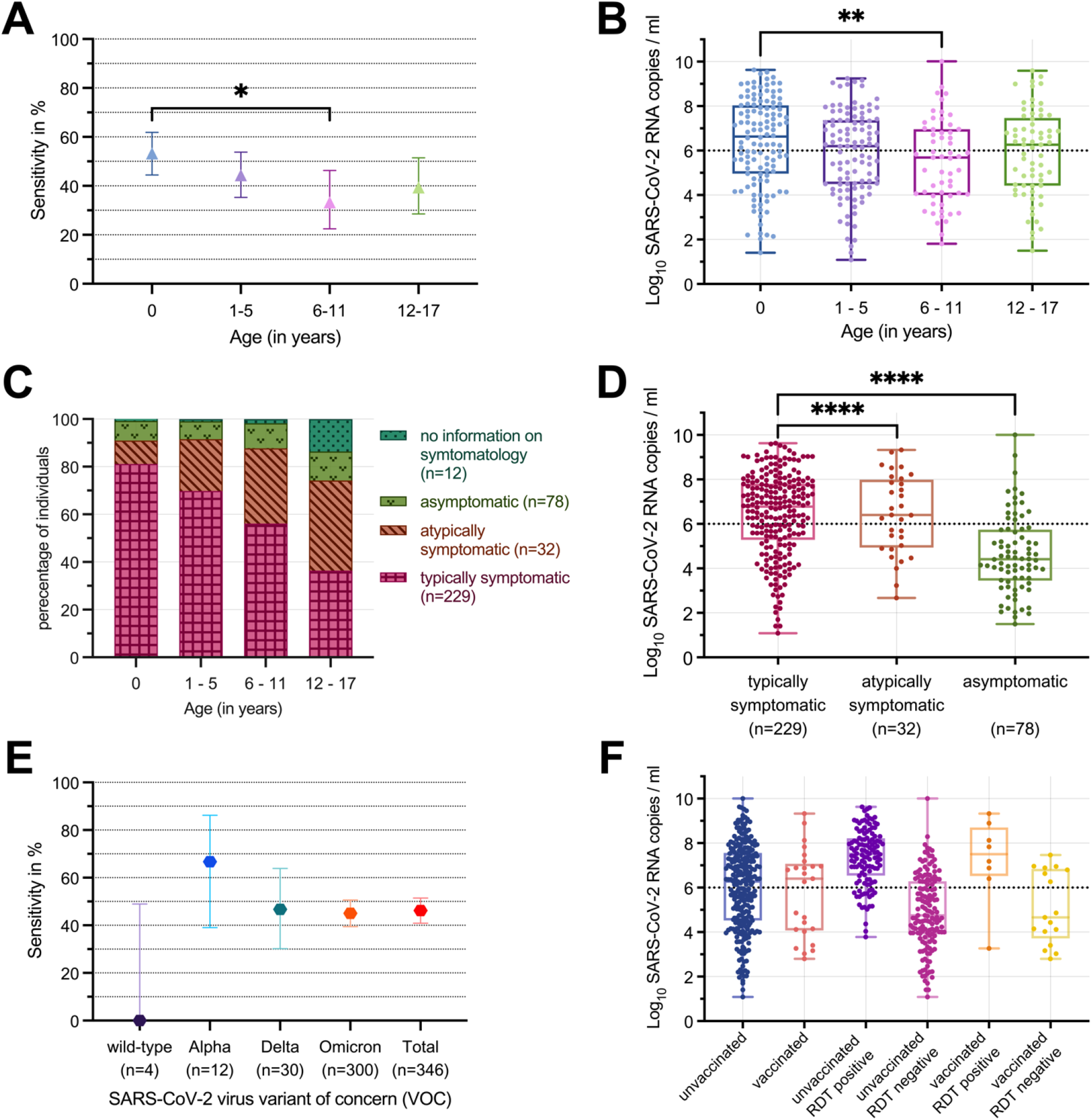
RDT performance in comparison to RT-qPCR stratified by age categories, VOC, and COVID-19 symptomatology **Figure 5A, 5B and 5C**: RDT sensitivity (n=351), logarithmised viral load (n=348), and symptomatology (n=351) in SARS-CoV-2 RNA copies/ml stratified by age categories (first year of life, 1 to 5 years, 6 to 11 years, 12 to 17 years). **Figure 5D** portrays the logarithmised viral load in SARS-CoV-2 RNA copies/ml, separated by RDT and COVID-19 symptomatology, n=339. The viral load of specimen of typically COVID-19 symptomatic children and adolescents exceeded statistically significant the viral load of atypically symptomatic or asymptomatic individuals. **Figure 5E** included 346 specimens with either molecularly confirmed or epidemiologically assigned VOC (in case of no molecular VOC diagnostics or, if available, known VOC of the infection source, VOC was assigned based on the VOC corresponding to at least 90% of the German COVID-19 cases at RDT performance). **Figure 5F:** Viral load in 306 specimens with known COVID-19 vaccination status stratified by vaccination status only and by vaccination status and RDT result. The viral load threshold of 10^6^ SARS-CoV-2 RNA copies/ml, suggested as infectivity threshold, is added as horizontal dotted line to Figure 5B, 5D and 5F.^25^ n: Number of enrolled RDT per group RDT: Antigen rapid diagnostic test RT-qPCR: Quantitative reverse transcription polymerase chain reaction *: p<0·05 **: p<0·01 ****: p<0·0001

### Relation to days since symptom onset

Among the 351 RT-qPCR positive tested samples of children and adolescents representing a COVID-19 typical symptomatology,^22^ in 177 cases the days since symptom (DSO) onset were retrieved. DSO ranged from 0 days (symptom onset on the day of RT-qPCR testing) up to 30 days (RDT 30 days since symptom onset; median: 1 DSO, IQR: 0-1 DSO). Maximum median viral load was detected on the first day since symptom onset (1·6×10^7^, IQR: 4·0×10^5^-1·1×10^8^ SARS-CoV-2 RNA copies per ml), followed by a median viral load of 6·8×10^6^ (IQR: 3·0×10^5^-1·3×10^8^) SARS-CoV-2 RNA copies per ml on the day of symptom onset with a following decreasing viral load.

### Influence of age on viral load, RDT performance and COVID-19 symptomatology

The performed 9,760 RDT were categorised by age: 1,566 (16·0%) children in the first year of life, 3,314 (34·0%) aged 1 to 5 years, 1,890 (19·4%) aged 6 to 11 years and 2,990 (30·6%) aged 12 to 17 years.

RDT sensitivity ranged from 52·3% (65/122, 95% CI 44·5%–61·9%) for children in the first year of life, to 44·3% (47/106, 95% CI 35·3%–54·0%) for the children aged 1 to 5 years, 33·3% (19/57, 95% CI 22·49%– 46·28%) for children aged 6 to 11 years and 39·4% (26/66, 95% CI 28·5%–51·5%) for children and adolescents aged 12 to 17 years (*Figure 5A*).

The following age-stratified median viral load was obtained: 4·3×10^6^ (IQR: 9·1×10^4^-1·1×10^8^) SARS-CoV-2 RNA copies per ml among children in the first year of life, 1·6×10^6^ (IQR: 3·1×10^4^-2·3×10^7^) SARS-CoV-2 RNA copies per ml among children aged 1 to 5 years, 5·0×10^5^ (IQR: 1·0×10^4^-9·1×10^6^) SARS-CoV-2 RNA copies per ml among children aged 6 to 11 years, 1·9×10^6^ (IQR: 2·6×10^4^-2·9×10^7^) SARS-CoV-2 RNA copies per ml among children and adolescents aged 12 to 17 years (*Figure 5B)*.

Children between 6 and 11 years showed a significantly reduced RDT sensitivity (p=0·016, Fisher’s exact test, *Figure 5A*) and viral load (p=0·0033, Mann-Whitney U test, *Figure 5B*) compared to children in the first year of life going in line with a lower rate of typically symptomatic children in this age group (*Figure 5C*).

The proportion of typically symptomatic individuals decreased with increasing age (*Figure 5C*).

### Prevalence of VOC

In total, 44 (14·2%) of the enrolled 351 RDT/RT-qPCR tandems underwent successful molecular VOC determination, 303 further samples were epidemiologically assigned to a VOC based on SARS-CoV-2 prevalence data.^19^ Consequently, in 346 RDT/RT-qPCR tandems the VOC was obtained: 1·2% (4/346) as wild-type SARS-CoV-2, 3·5% (12/346) as Alpha VOC, 8·7% (30/346) as Delta VOC, and 86·7% (300/346) as Omicron VOC. 4 specimens remained without VOC assignment due to prevalence levels of any VOC below 90% at sampling time. One further specimen was not considered due to a result of variant-specific PCR testing and spike protein sequencing which did not match any commonly defined VOC.

Due to the low sample size for wild-type SARS-CoV-2, no valid VOC stratified sensitivity data could be determined (0·0%, 0/4, 95% CI 0·0%–49·0%). RDT sensitivity was higher in Alpha VOC (66·7%, 8/12, 95% CI: 39·1%–86·2%) compared to Delta VOC (46·7%, 14/30, 95% CI 30·2%–63·9%), and Omicron VOC (45·0%, 135/300, 95% CI 39·5%–50·7%). Differences in VOC specific sensitivity were not significant (pairwise comparisons using Fisher’s exact test, all p>0·08, *Figure 5E*).

### Influence of COVID-19 vaccination on RDT performance

In 7,990 of 9,760 (81·9%) enrolled RDT information on COVID-19 vaccination status at RDT performance was available: 6,977 RDT (87·3%) were conducted on unvaccinated children and adolescents, 1,013 (12·7%) on individuals who had received at least one dose of COVID-19 vaccine.

Among unvaccinated children and adolescents, RDT sensitivity was 44·8% (126/281, 95% CI 39·1%– 50·7%), RDT specificity 99·8% (6,681/6,696, 95% CI 99·6%–99·9%). Among vaccinated participants, RDT sensitivity was 32·00% (8/25, 95% CI 17·2%–51·6%), specificity 100·00% (988/988, 95% CI 99·6%–100·0%). Differences in sensitivity (p=0·29) and specificity (p=0·24, both Fisher’s exact test) were not significant.

Viral load did not significantly differ comparing COVID-19 unvaccinated (median viral load 1·9×10^6^, IQR: 3·3×10^4^-3·6×10^7^ SARS-CoV-2 RNA copies per ml) with vaccinated children and adolescents (median viral load 2·5×10^6^, IQR: 1·2×10^4^-1·2×10^7^ SARS-CoV-2 RNA copies per ml, p=0·20, Mann-Whitney U test, *Figure 5F*).

Of the 281 specimens on unvaccinated participants, 278 could be allocated to a VOC: 1 (0·4%) wild-type SARS-CoV-2, 8 (2·9%) Alpha VOC, 28 (10·0%) Delta VOC, and 241 (86·1%) Omicron VOC containing samples. All 25 vaccinated SARS-CoV-2 positive children and adolescents were assigned to Omicron VOC. Considering only Omicron VOC infections, RDT sensitivity was slightly higher for unvaccinated (110/241, 45·64%, 95% CI 39·5%–52·0%) compared to vaccinated (8/25, 32·00%, 95% CI 17·2%– 51·6%) children and adolescents without any statistical significance (p=0·21, Fisher’s exact test).

## Discussion

The obtained overall sensitivity of 44·7% in the cohort of children and adolescents corresponds to a previously published laboratory assessment without consideration of age and therewith connected influencing factors (36·0% to 64·0%).^27^ The World Health Organization’s (WHO) recommendations for RDT level of accuracy of ≥97% specificity were achieved, but the claimed minimum RDT sensitivity of 80·0% was missed.^28^ With respect to the poorly studied RDT accuracy in children and adolescents so far, the presented performance parameters scores are at the lower end. However, they were obtained in a study which resulted in more case numbers in a real-life point-of-care setting than in previous publications.9,10,12,13,17

The described difference may be explained as a possible consequence of including asymptomatic individuals and screening the entire hospital instead of only considering the paediatric emergency department and symptomatic patients.^9^ The data presented is the first to analyse RDT performance in a paediatric cohort in the context of the current spreading Omicron VOC as well as a significant subset of COVID-19 vaccinated children and adolescents. In the data presented, no statistically significant differences could be observed in RDT performance between different VOC.

Reliability of RDT performance clearly depended on specimens’ viral load which is influenced by age and days since symptom onset (DSO). The differences in the age-stratified viral load levels may be explained by the different proportions of COVID-19 symptomatology in the several age categories: With increasing age, the trend of an ascending proportion of atypically and asymptomatic individuals accompanied by a descending viral load was obtained. Consequently, the age-depended differences in RDT performance can be explained in differences of the study population. Within the clinical framework, the different reasons of hospitalisation in the several age categories may have led to this characterisation: As children in the first year of life with typical COVID-19 symptomatology may enlist the hospitals’ medical care early and large-scale, the school children aged 6 to 11 years may be detected coincidentally in the COVID-19 screening in the context of a differing reason for medical consultation.

In contrast to previous evidence,^18^ no significantly reduced viral load was observed in Omicron VOC. This may be explained by the dominating proportion of Omicron VOC in this study. RDT sensitivity did not correlate with immunisation status which has been reported in a preprint analysis as factor impairing RDT sensitivity among vaccinated individuals.^14^

The study is limited in several aspects: Data collection in the real-life and point-of-care setting led to differing distributions and proportional use of the three RDT across the contributing clinical departments and over the temporal course. As each child and adolescent was tested by only one of the three used RDT, the direct comparability between the manufacturers’ is limited. Comparison of VOC was limited as case numbers for pre-Omicron VOC cases were low compared to Omicron VOC which was detected in most SARS-CoV-2 infections. Specimen collection for both diagnostics, RDT and RT-qPCR, was carried out by professional operators. The potential influence and inhomogeneity in sampling, especially in consideration of the preanalytical challenges in children, in test execution as well as in interpretation is probable. Molecularly based VOC determination was only carried out on RT-qPCR samples from January 2021 to January 2022 when it was stopped due to high incidence levels and dominance of the Omicron VOC in the pandemic course.^4,19^ Due on this, for a relevant share of the study-included SARS-CoV-2 positive samples, the VOC determination was deduced as an epidemiological assignment. The proportion of RDT on COVID-19 vaccinated children and adolescents among the entire SARS-CoV-2 positive cases is reduced and requires further evaluations with an extended cohort. Despite these limitations, the study presents an analysis of real-life data in a large population of children over an extended study period covering several VOC.

For children and adolescents, the indication, as well as advantages and disadvantages for RDT usage is comparable to the one for adults.^18,20^ If no RT-qPCR is available, positive RDT results during high incidence periods can be assumed as SARS-CoV-2 positive. In symptomatic children and adolescents, a repetition of a negative RDT on the next day may probably increase diagnostic reliability. Due to the low sensitivity in asymptomatic individuals, the usefulness of RDT seems limited in large-scale SARS-CoV-2 screening programs. This intrahospital assessed data on RDT reliability should also be considered in terms of RDT screening usage, including its self-testing option for children and adolescents as COVID-19 management strategy in the context of schools and nurseries.^29,30^

## Data Availability

Individual participant data that underlie the results reported in this article after deidentification is available on request immediately following publication ending 5 years following article publication to researchers who provide a methodological sound proposal to achieve aims in the approved proposal. Proposals should be directed to.

## Data Access, Responsibility, and Analysis

Ms Wagenhäuser and Dr Krone had full access to all the data in the study and take responsibility for the integrity of the data and the accuracy of the data analysis.

## Authors’ contributions

Conception and design: Krone, Forster, Weißbrich, Dölken, Kurzai, Vogel, Härtel, Liese, Andres.

RT-qPCR testing, standardised Ct quantification, and VOC determination: Knies, Hofmann, Weißbrich, Dölken.

RDT use and documentation instruction: Krone, Taurines, Flemming, Meyer, Böhm, Scherzad, Liese, Andres.

User support: Krone, Eisenmann, Rauschenberger, Vogel, Liese, Andres.

Collection of clinical data from patient’s files: Krone, Wagenhäuser, Engels, McDonogh. Statistical analysis: Krone, Wagenhäuser, Gabel, Petri, Reusch.

Obtained funding: Dölken, Kurzai, Vogel.

First draft of the manuscript: Krone, Wagenhäuser.

The manuscript was reviewed and approved by all authors except Professor Ulrich Vogel.

Professor Ulrich Vogel had a major contribution to the concept and design of the study as well as in obtaining funding, supervising and supporting the users. As he passed away during manuscript drafting, he was not able to review and approve the final version of the manuscript. We miss him as an enthusiastic college and friend who showed a great dedication to his work, family and friends.

## Additional contributions

We thank all hospital staff for conducting RDT testing and documenting test results and all laboratory staff in the virological diagnostic laboratory for performing RT-qPCR testing. We thank accounting department from medical controlling for SAP support.

## Information on previous presentation of the information

1,034 RDT results on children and adolescents have already been included in an age independent RDT performance assessment in a cohort of 5,068 RDT with data collection from the 12^th^ of November 2020 to the 28^th^ of February 2021^20^ as wells as the sequel up to the 30^th^ of January 2022 including 35,479 specimen within 5,623 on children and adolescents.^18^

## Acknowledgments

This study was funded by the Federal Ministry for Education and Science (BMBF) via a grant provided to the University Hospital of Wuerzburg by the Network University Medicine on COVID-19 (B-FAST, grant-No 01KX2021), by the Bavarian Staten Ministry of Health and Care via Bay-VOC as well as by the Free State of Bavaria with COVID-research funds provided to the University of Wuerzburg, Germany.

Nils Petri is supported by the German Research Foundation (DFG) funded scholarship UNION CVD.

